# Protocol for the British Paediatric Surveillance Study of Neonatal Stroke in the United Kingdom and the Republic of Ireland in babies in the first 90 days of life

**DOI:** 10.1101/2022.04.01.22273284

**Authors:** T’ng Chang Kwok, Robert A Dineen, William Whitehouse, Richard M Lynn, Niamh McSweeney, Don Sharkey

**Affiliations:** Centre for Perinatal Research, School of Medicine, University of Nottingham, E floor, East Block, Queen’s Medical Centre, Nottingham, NG7 2UH; Radiological Sciences, Mental Health & Clinical Neuroscience, School of Medicine, University of Nottingham, Queen’s Medical Centre, Nottingham, NG7 2UH; NIHR Nottingham Biomedical Research Centre, Nottingham; Population, Policy and Practice Research and Teaching Department, University College London Great Ormond Street Institute of Child Health, 30 Guilford Street, London, WC1N 1EH; Department of Paediatrics and Child Health, Cork University Hospital, Wilton, Cork, T12 DC4A, Ireland

**Keywords:** Infant, Neonatal Stroke, Population Surveillance

## Abstract

**Background:** Neonatal stroke is a devastating condition that causes brain injury in babies and often leads to lifelong neurological impairment. Recent, prospective whole population studies of neonatal stroke are lacking. Neonatal strokes are different from those seen in older children and adults. A better understanding of the aetiology, current management and outcomes of neonatal stroke could reduce the burden of this rare condition. Most healthcare professionals see only a few cases of neonatal stroke in their careers, so population-based prospective studies are needed.

**Objectives:** To explore the incidence and two-year outcomes of neonatal stroke across an entire population in the UK and the Republic of Ireland.

**Population:** Any infant presenting with neonatal stroke in the first 90 days of life.

**Design:** Active national surveillance study using a purpose-built integrated case notification-data collection online platform.

**Methods:** Over a 13-month period, British and Irish clinicians will notify any cases of neonatal stroke electronically via the online platform monthly. Clinicians will complete a primary questionnaire via the platform detailing clinical information, demographic details and investigations, including neuroimaging for detailed analysis and classification. An outcome questionnaire will be sent at two years of age via the platform. Appropriate ethical and regulatory approvals have been received from England, Wales, Scotland, Northern Ireland and the Republic of Ireland.

**Conclusion:** The neonatal stroke study represents the first multinational population surveillance study delivered via a purpose-built integrated case notification-data collection online platform and data safe haven, overcoming the challenges of setting up the study.

**Synopsis:** *Study question:* The neonatal stroke active surveillance study aims to explore the incidence and two-year outcome of neonatal stroke in the UK and Ireland.

*What is already known?:* Neonatal stroke is a rare but often devastating condition with lifelong consequences including cerebral palsy, epilepsy and cognitive delay. There are no contemporary, prospective multinational population studies on the presentation and outcomes of neonatal stroke. Whilst often the aetiology is multifactorial further information on underlying aetiology may help to identify potential future preventative treatments leading to improved outcomes.

*What does this study add?:* International collaboration is required to understand the epidemiology, management and outcomes of rare diseases or conditions. This is the first multinational surveillance study delivered via a purpose-built integrated case notification-data collection online platform and data safe haven, presenting practical and ethical challenges. The study will describe the burden of neonatal stroke while providing parents/carers and healthcare professionals with up-to-date information about the condition including the two-year outcomes.

## Background

Neonatal stroke is a devastating condition that causes brain injury and often leads to lifelong neurological impairment^1^ including cerebral palsy^2, 3^ and global developmental delay in particular language and cognitive delay ^4, 5^. A common presenting symptom is focal seizures in the neonatal period with a later incidence of 15-25% of epilepsy depending on the extent and location of the injury^6^. Infants with neonatal stroke will continue to depend on healthcare services throughout childhood. It is estimated that each case of neonatal stroke will cost the healthcare services at least £26,000 (∼31,000 EUR; 34,000 USD) for the initial admission and a further £40,000 (∼48,000 EUR; 52,000 USD) during the first five years of life^7^. There will also be an increasing need for education services to support survivors of neonatal stroke^8^. The impact on family life and loss of economic productivity due to neonatal stroke is also known to be significant^9, 10^.

Neonatal strokes are different from strokes seen in older children and adults^11^. In babies, the brain is immature with greater capacity for plasticity in response to brain injury and hence greater potential for recovery^12^. However, the susceptibility of the developing newborn brain may make it more prone to injury leading to long-term impairment compared to older children. A better understanding of neonatal stroke will guide further research in identifying new interventions to prevent and treat neonatal stroke^13^.

Although stroke is more common in the first 28 days of life than in older children^14^, the true contemporary incidence of neonatal stroke is still unclear. Previous Canadian^14^ and UK studies^15^ from over 20 years ago reported the incidence of neonatal stroke between 5.2 – 10.2 per 100,000 live births whereas a retrospective case-control study using neuroimaging^16^ found an incidence of 37 per 100,000 live births. Previous retrospective case-control studies^17, 18^ were based on infants born in a province or small geographical area, with data collected over 20 years ago, limiting the generalisability of the findings.

As most clinicians may see only a few cases of neonatal stroke in their career, there is a variation in practice and guidance in the management of neonatal stroke^19^. This is unlike stroke in adults where much more is known. Hence, there is a need for an active surveillance study with follow-up to have a better understanding of the condition. The challenges of setting up a multinational electronic surveillance system as well as the regulatory approval process across multiple nations will also be discussed.

## Aims

This active surveillance study aims to explore the burden of neonatal stroke in the UK and the Republic of Ireland (ROI) to better allocate resources and improve the care of babies with neonatal stroke.

### Primary Objectives

1. To estimate the incidence of stroke presenting in infants in the first 90 days of life in the UK and the ROI.
2. To determine the two-year outcome of neonatal stroke, including neurodevelopmental outcome.

### Secondary Objectives

1. To analyse the proportion of the different types of neonatal stroke (arterial ischaemia, venous thrombosis, haemorrhage) and their impact on presentation and outcome
2. To analyse known predisposing maternal and infant factors for neonatal stroke
3. To analyse the presentation of infants with neonatal stroke
4. To analyse the variation in clinical investigations used in neonatal stroke
5. To describe findings of investigations, particularly neuroimaging
6. To analyse the variation in management of neonatal stroke including the follow-up pathway

## Methods

### Study design

The study represents the first multinational population surveillance study using the British Paediatric Surveillance Unit (BPSU) active surveillance approach delivered via a purpose-built integrated case notification-data collection online platform and data safe haven, developed by BPSU in collaboration with the University of Dundee Health Informatics Centre (HIC) (**Figure 1**).

**Figure 1:**
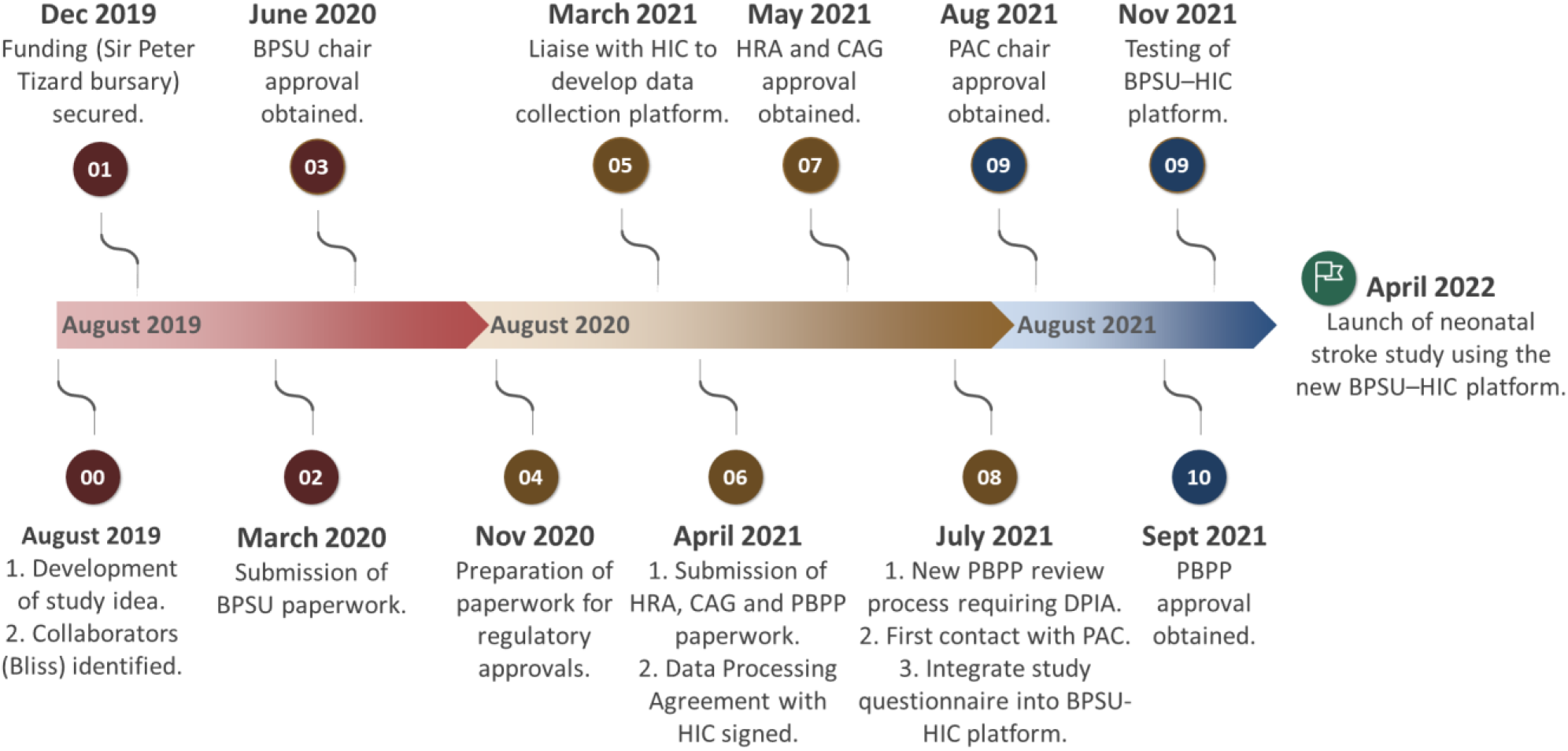
Timeline depicting the launch of the neonatal stroke surveillance study from the conceptualisation of the research idea. Study progress was halted for a year in March 2020 due to the COVID-19 pandemic. BPSU = British Paediatric Surveillance Unit. HIC = Health Informatics Centre. HRA = Health Research Authority. CAG = Confidentiality Advisory Group. PBPP = Public Benefit and Privacy Panel. DPIA = Data Protection Impact Assessment. PAC = Privacy Advisory Committee of Northern Ireland.

### Active surveillance

Over a 13-month period, anonymised notifications of the stroke cases seen presenting in babies in the first 90 days of life will be provided to the BPSU using the BPSU Orange e-Card delivered via the BPSU–HIC platform. The BPSU Orange e-Card has a list of rare disorders under active surveillance which is sent electronically every month to over 4,200 British and Irish consultant paediatricians and other specialities. Information leaflets and study newsfeeds will be circulated to all clinicians receiving the BPSU Orange e-Card during the study period.

Clinicians will indicate any cases of stroke seen by them in babies up to 90 days of age that meet the case definition, or ‘nothing to report’ on the monthly BPSU e-orange card system and a response is expected. The ‘active surveillance’ is an important feature of the surveillance scheme as it allows the compliance of the system to be continually monitored, ensuring good coverage of the paediatric surveillance scheme across the UK and ROI^20^. The reporting compliance rate in 2019 was 90% ensuring that the proposed study methodology will have high case ascertainment, validity and reliability.

The notifying clinician will then be directed to an online initial questionnaire (**Appendix 1**) via the BPSU–HIC platform and complete the electronic questionnaire. The initial questionnaire (**Appendix 1**) will seek information about the presentation of neonatal stroke, demographic details, investigations including neuroimaging, and treatment received. On completion of the initial questionnaire, the reporting clinicians will be asked to retain the patient details for a follow-up outcome questionnaire, which will be sent at two years of age (**Appendix 2**) through the BPSU–HIC platform (**Figure 2**). Each questionnaire will be date- and time-stamped on completion and specific to the notifying clinician. Separate questionnaires will be generated for Northern Ireland clinicians (**Appendix 3 and 4**) due to differences in regulatory requirements.

**Figure 2:**
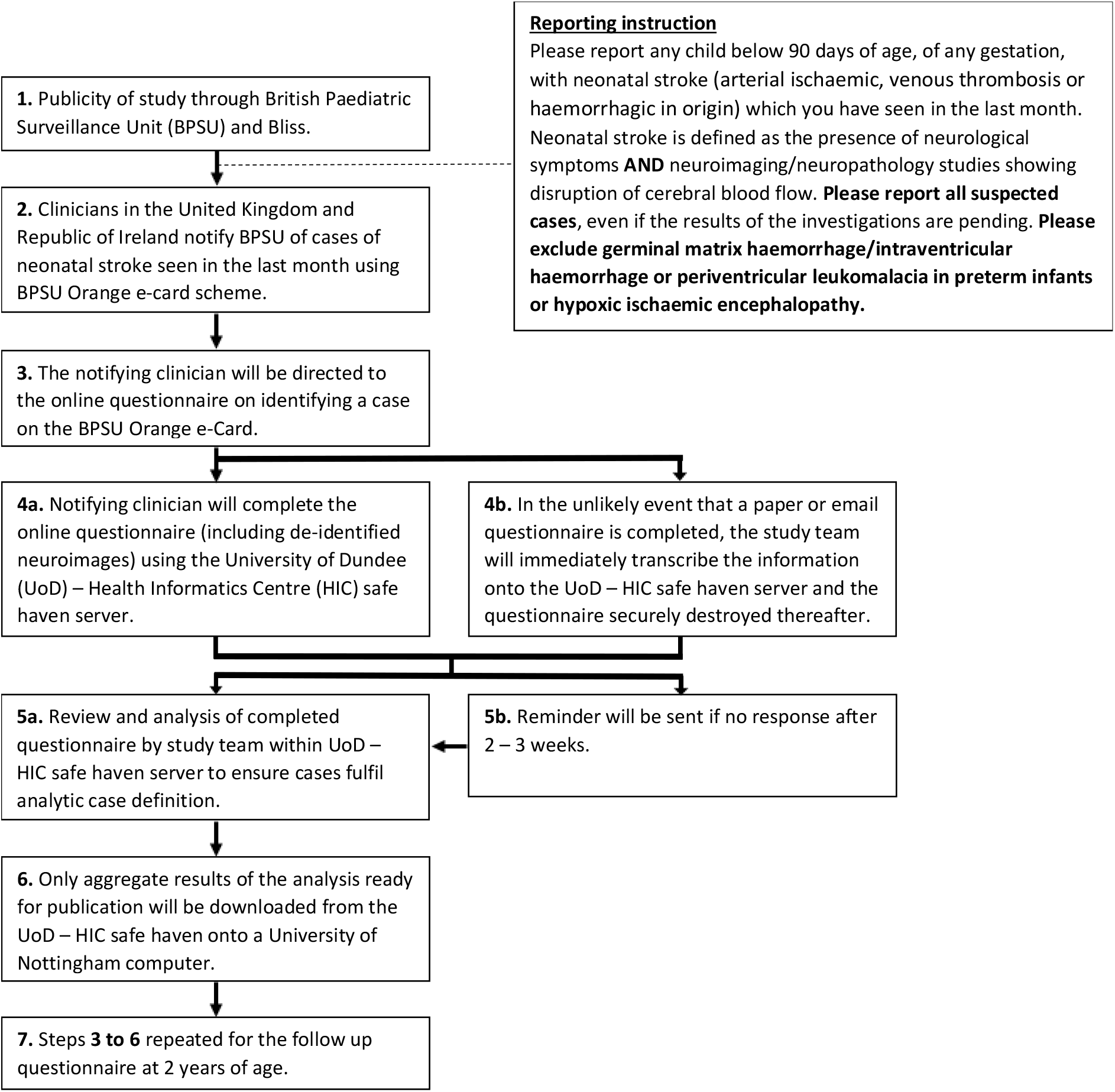
Flow diagram depicting the notification and data flow during the neonatal stroke surveillance study.

### Case definition

#### Inclusion criteria

Any infant from birth till 90 days of age, of any gestation, with neonatal stoke defined as:-

1. Developing any neurological symptoms (including seizure, neurological deficit, lethargy, abnormal tone, poor feeding) **AND fulfilment of either 2a OR 2b:**
2a. Neuroimaging (such as MRI or CT) showing disruption or evidence of disruption of cerebral blood flow. The radiology findings include either:
  i. Arterial ischaemia: defined as the presence of: (i) partial or complete occlusion of the cerebral artery(ies) in relation to a focal brain lesion; or (ii) brain lesion pattern with imaging that can only be explained by occlusion of a specific cerebral artery(ies); **OR**
  ii. Venous thrombosis: defined as the presence of a thrombus within the cerebral vein(s) or venous sinus(es), with partial or complete occlusion. **OR**
  iii. Haemorrhage: defined as the presence of a haemorrhage in the parenchyma of the brain.
2b. Neuropathologic studies (such as post-mortem) showing disruption or evidence of disruption of cerebral blood flow

#### Exclusion criteria

1. Hypoxic ischaemic encephalopathy
2. Germinal matrix haemorrhage/intraventricular haemorrhage or periventricular leukomalacia in preterm infants (defined as below 37 weeks of gestational age)
3. Metabolic injury such as kernicterus and hypoglycaemia
4. Encephalitis (bacterial and viral)
5. Accidental or non-accidental injury
6. Vascular anomalies such as arteriovenous malformation
7. Brain tumour

The eligibility criteria above are based on the 2006 international National Institute of Child Health and Human Development (NICHD) and the National Institute of Neurological Disorders and Stroke (NINDS) workshop^21^ which defined perinatal stroke as “a group of heterogeneous conditions in which there is focal disruption of cerebral blood flow secondary to arterial or cerebral venous thrombosis or embolization, between 20 weeks of fetal life through the 28th postnatal day, confirmed by neuroimaging or neuropathologic studies”. The radiological definition is based on Sebire et al^22^ used in the International Paediatric Stroke Study^2^ and Govaert et al^23^. The exclusion criteria will ensure that the study finding is comparable to other reports published nationally^24^ and internationally^23^.

As this study is focused on neonatal stroke rather than fetal stroke, the age definition for the study is from birth. The age range extends till 90 days of age to capture infants that present late with either neuroimaging or neuropathologic studies, but which suggested stroke occurring in the perinatal period. The number of cases between 28 and 90 days of age is expected to be minimal^14^. As we cannot be completely certain that the neuroimaging or neuropathologic studies accurately date the neonatal stroke to have occurred in the first 28 days, we will separate the cases of neonatal stroke presenting within 28 days and those presenting between 28 to 90 days.

### Data collection

The online questionnaire was developed and delivered using the BPSU–HIC platform. Data entered by notifying clinicians will be stored within the HIC safe haven server which is ISO 270001 standard certified (https://www.dundee.ac.uk/hic/hicsafehaven/). If clinicians are unable to use the online system, questionnaires will be sent out in electronic format to the reporting clinicians via a secure email account (ISO 9001 Standards certified). As a last resort, a paper questionnaire form will be sent. It will be marked confidential and double enveloped for data collection with a self-addressed envelope to be returned to the research team via registered mail. In the unlikely event of email or paper questionnaires being completed, the study team will immediately transcribe the information onto the HIC safe haven server on behalf of the notifying clinicians and the questionnaires destroyed thereafter.

All the requested information will be obtained using data recorded in case notes and laboratory/radiology results only, without further direct contact or active participation with patients. No extra investigations will be sought. De-identified images from neuroimaging studies will be uploaded by the notifying clinicians onto the HIC safe haven server where possible. Images can also be sent using an encrypted CD, which are then uploaded on the HIC safe haven server immediately and the CDs destroyed by the research team. If the images are not available, a copy of the de-identified neuroimaging report will be requested.

Limited patient identifiable data will be obtained for study validity reasons to identify duplicate cases and track follow-up. Each case will be pseudonymised through a unique computer-generated identifier number. The minimal identifiable data collected for study validity purposes will be removed as soon as it is practically possible and will be kept separate from the rest of the anonymised clinical data linked by the unique identifier number. All patient identifiable data collected for identifying duplicate cases will be destroyed once the data collection process is complete.

All data will be collected using opt-out consent. Public information leaflets and flyers will be disseminated to participating clinicians and on the BPSU website. These will provide information about the neonatal stroke study as well as how families can opt-out of babies’ medical information from being used for research by informing their attending clinician. In the unlikely event that families decide to opt-out, a small amount of identifiable information will be held for the duration of the study to ensure that no further clinical information is collected.

### Data management and security

All study data will be collected, analysed and held securely on the HIC safe haven server, with access restricted by user login identifiers and passwords, which are encrypted using a one-way encryption method. The HIC safe haven server is ISO 270001 certified and is externally audited twice yearly to ensure compliance and is overseen by the HIC Information Governance Committee, with National Health Service (NHS) and legal membership. Only aggregate results of the analysis ready for publication will be downloaded from the HIC safe haven server. In keeping with standard practice^25^, all anonymised data will be archived within HIC safe haven server for 20 years.

### Analysis

Primary analyses will be based on all cases that meet the analytic case definition. The patterns of availability of data and timing of data collection will be summarised, including differentiation of fully, or partially completed variables from those completely missing. The aggregation of data variables will be described. Descriptive statistics will be presented as numbers and percentages with 95% confidence intervals (CI) estimated using the binomial exact method, for binary and categorical variables; means with standard deviations for normally distributed continuous variables; and medians with interquartile range for skewed continuous variables. Neuroimages collected will be reviewed by paediatric neuroradiologists within the research team (RD) to identify concordance with the reported findings entered by reporting clinicians.

Estimation of incidence will include the first month of surveillance based on the date of diagnosis. Incidence rates (IR) will be estimated for the whole sample and by gender. Birth statistics used for the denominators for incidence estimation will be obtained by gender from:

- Office for National Statistics (ONS; England and Wales).
- National Records of Scotland (NRS; Scotland).
- Northern Ireland Statistics and Research Agency (NISRA; Northern Ireland).
- Central Statistics Office (CSO; Republic of Ireland)

### Patient and public involvement

The neonatal stroke surveillance study was designed in partnership with Bliss, the national charity for families of babies born sick or premature as well as lay representatives of the BPSU scientific committee. Bliss is supportive of the research question and the need to collect information about neonatal stroke to improve the support and care provided to the babies and families affected.

### Regulatory approval

The study had received ethical approval from the Nottingham 1 Research Ethics Committee (Reference 21/EM/0110) supported by the Health Research Authority on advice from the Confidentiality Advisory Group (England and Wales) (Reference 21/CAG/0061); the Public Benefit and Privacy Panel (Scotland) (Reference 2122-0006 Kwok); and the chair of the Privacy Advisory Committee (Northern Ireland).

### Dissemination

The study is in the process of being registered with ClinicalTrials.gov. Study findings will be disseminated to the scientific community via peer-reviewed publications and conference presentations. The research team will also disseminate the findings to clinicians, parents/carers and the public via the partnership with Bliss, British Association of Perinatal Medicine and British Paediatric Neurology Association. Anonymised aggregate data will be made available at the time of peer-reviewed scientific publications either via online supplementary material or the University of Nottingham research data repository.

### Comment

#### Sample size

It is estimated that our study will detect 150 cases of neonatal stroke per year. The Canadian paediatric stroke registry^14^ reported the incidence of neonatal arterial ischaemic stroke to be 10.2 per 100,000 live births between January 1992 to December 2001 while the 2001 UK surveillance study^15^ identified 31 neonatal stroke cases annually (5.2 per 100,000 live births^26^). Both studies^14, 15^ collected data over 20 years ago and when diagnostic imaging and reporting of neonatal stroke was not as advanced. The recent retrospective population-based cohort study using the national neonatal electronic database^24, 27^ reported a neonatal stroke incidence of 11 – 15 per 100,000 live births in England between 2012 to 2017. However, the study^24, 27^ excluded babies cared for outside the neonatal unit and only contains basic information on babies, without distinguishing between the different types of neonatal stroke and two-year outcome. A population-based birth prevalence of disease-specific perinatal stroke in Alberta by Dunbar et al^28^ included prospective data collection between 2008 and 2017 and retrospective data between 1990 and 2008. They reported an overall prevalence of neonatal stroke of 1 in 1,100 of which 1 in 3,000 was neonatal arterial ischemic stroke (NAIS), 1 in 7,900 for presumed NAIS, 1 in 6,000 for periventricular venous infarction, 1 in 9,100 for cerebral sinovenous thrombosis, and 1 in 6,800 for neonatal haemorrhagic stroke^28^.

#### Opt-out consent

The collection of a minimum dataset of identifiable patient information using opt-out consent is part of the conventional active surveillance methodology used by BPSU to prevent case ascertainment bias in the surveillance of a rare condition. Minimal identifiers are required to match duplicate cases reported to the study. If clinicians do not use the same reference number system, then more than one identifier, such as date of birth and gender, might be needed for matching. Some identifiers are also important pieces of clinical data in newborn infants, for example, the exact date of birth is required to calculate the age at diagnosis in days or weeks.

The incidence of neonatal stroke can only be estimated if information about all cases which occur within a specified period can be collected. As neonatal stroke is a rare condition, failure to obtain consent from only a few patients would make the incidence estimates inaccurate and under-ascertain cases. This may lead to incorrect conclusions being drawn, especially if the failure to obtain consent and data occurred in a single region or in babies with a less severe form of neonatal stroke. Furthermore, opt-out consent was found to be an acceptable method by clinicians and parents/carers in neonatal studies, including clinical trials^29^.

#### Collaborative standardised regulatory approval

Section 251 of the National Health Service (NHS) Act 2006 in the UK was established to allow confidential patient information to be used for essential NHS activities and important medical research where the use of anonymised information is not possible and seeking consent was not practical^30^. Hence, careful regulation of this process is needed to protect the interest of the patients and the public while facilitating appropriate use of the information. At present, England and Wales, Scotland, as well as Northern Ireland, have different pathways to seeking regulatory approvals for the use of patient information. A collaborative and standardised approach is needed across the four nations in the UK to avoid research wastage in completing different regulatory paperwork used in each nation while ensuring the research is carried out to the highest standard in minimising the disclosure risk while protecting confidentiality and interest of the patients and public.

#### Study impact

##### Clinical practice

This study will raise awareness of neonatal stroke among healthcare professionals looking after babies. This is important as most healthcare professionals will see only a few cases of neonatal stroke in their career, leading to potential delay in recognising the often-subtle initial signs of neonatal stroke. This study will also explore the variation of practice across the UK and Ireland to standardise the investigation and management of neonatal stroke.

##### Patients and families

This study will provide families caring for babies with neonatal stroke with up-to-date information about the condition including outcome at 2 years of age. This will consolidate information given by healthcare professionals and alleviate some anxiety families may have when first being informed of the diagnosis and caring for babies with neonatal stroke.

##### Public health

This study will estimate the burden of neonatal stroke. Healthcare professionals and policy developers will be able to use this information to better allocate resources in supporting babies with neonatal stroke.

##### Scientific community

Neonatal stroke is a distinct entity from childhood or adult stroke. It is multifactorial due to an interplay between maternal and fetal circulation as well as postnatal factors. There is much to learn on both the mechanism of injury and recovery^31^. Earlier detection may improve outcomes. This study will provide further insight into the epidemiology of neonatal stroke and help inform future studies looking at prevention and treatment options. From a surveillance study methodological point of view, the study will evaluate the feasibility and success of using a dedicated online platform for the collection of data on rare paediatric conditions.

## Data Availability

All data produced in the present study are available upon reasonable request to the authors.

## Acknowledgements

We would like to thank our study partners Bliss and BPSU for their support in designing the study as well as for publicising the study alongside the British Association of Perinatal Medicine and the British Paediatric Neurology Association. We are extremely grateful for the support from the Health Informatics Centre (University of Dundee) in developing the online platform used in the delivery of our questionnaires. We would also like to thank all researchers and clinicians who have provided feedback to the BPSU–HIC online platform. Full details of the study could be found on the BPSU website (https://www.rcpch.ac.uk/work-we-do/bpsu/neonatal-stroke).

## Appendices

### Appendix 1: Initial questionnaire

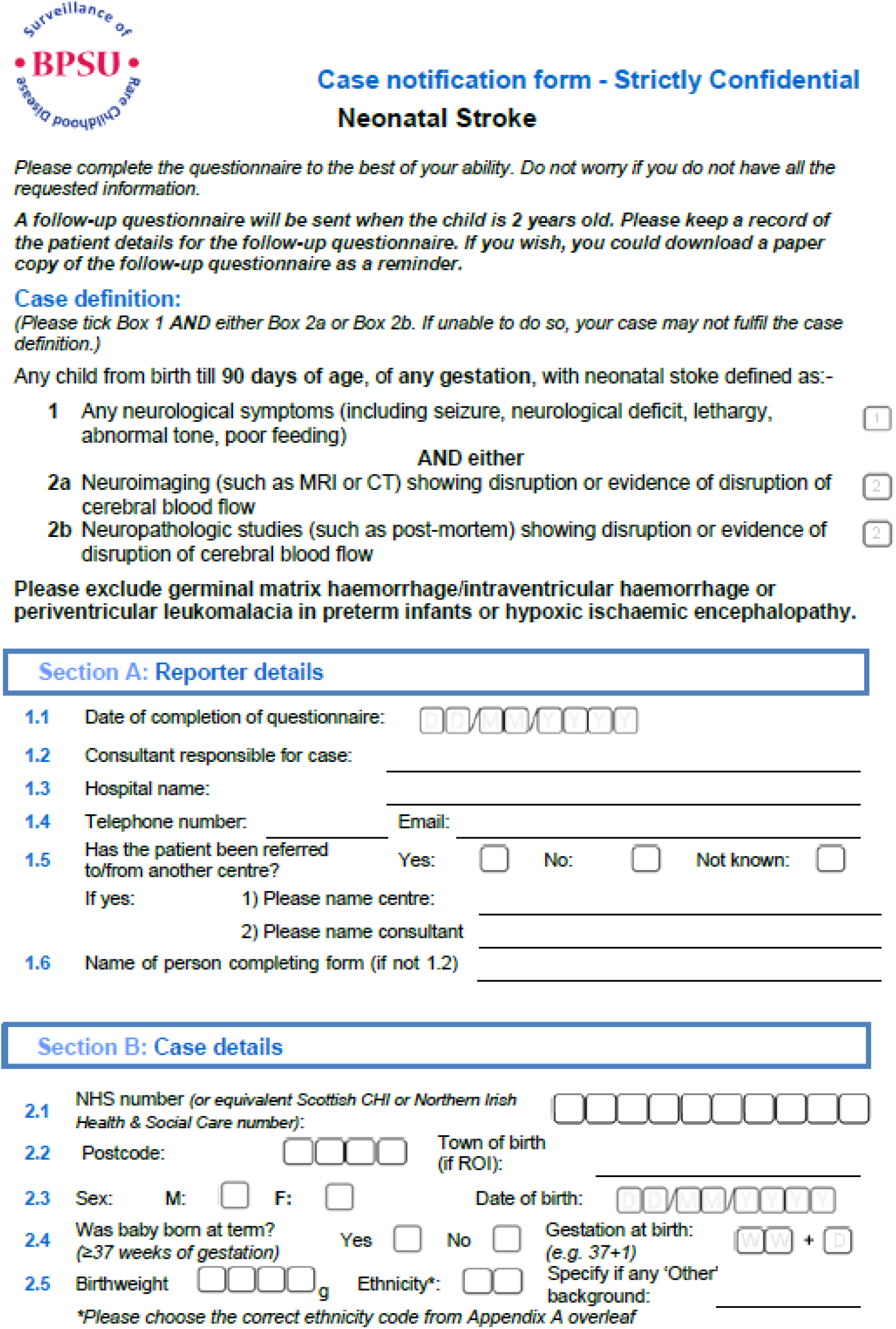
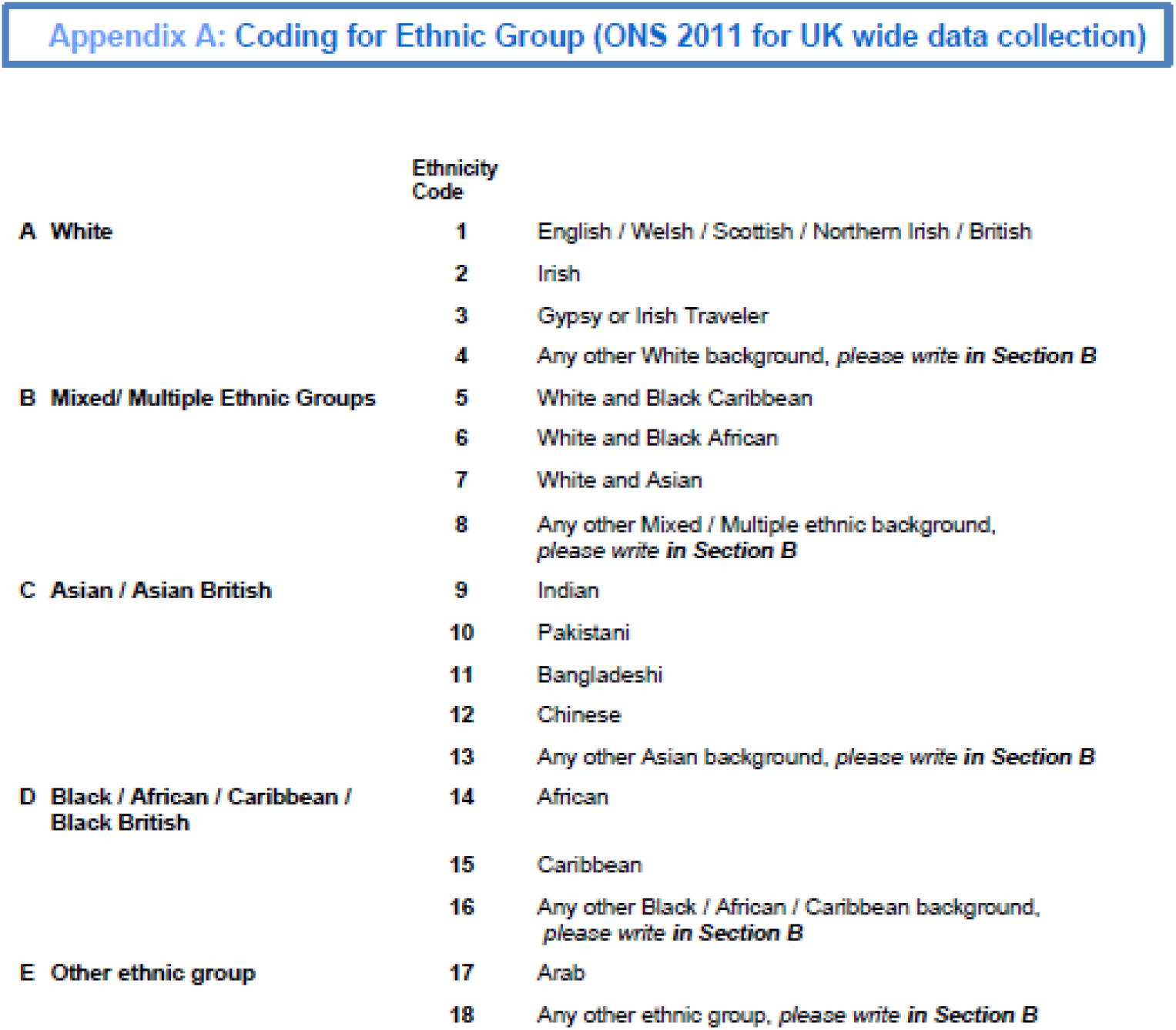
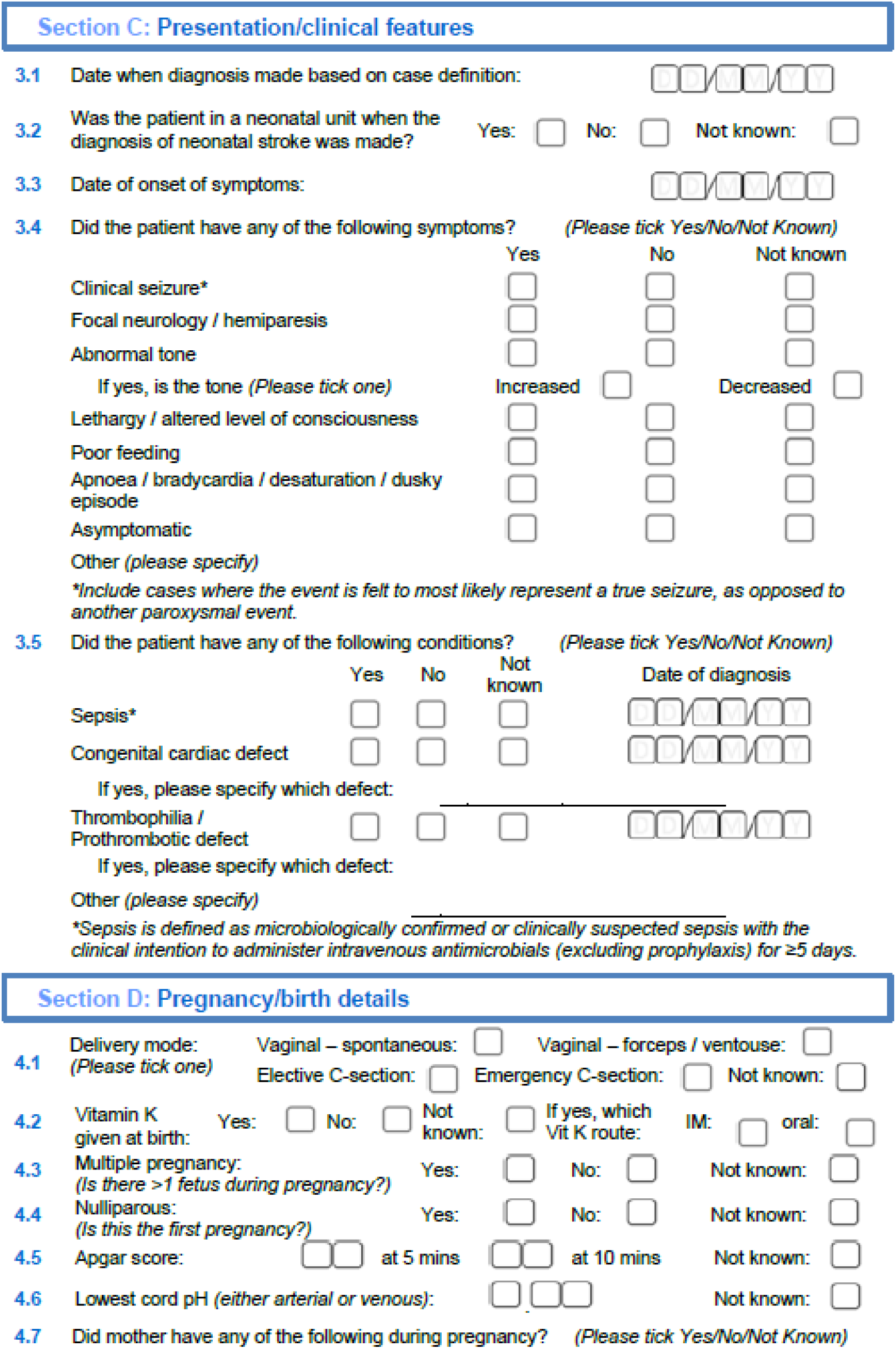
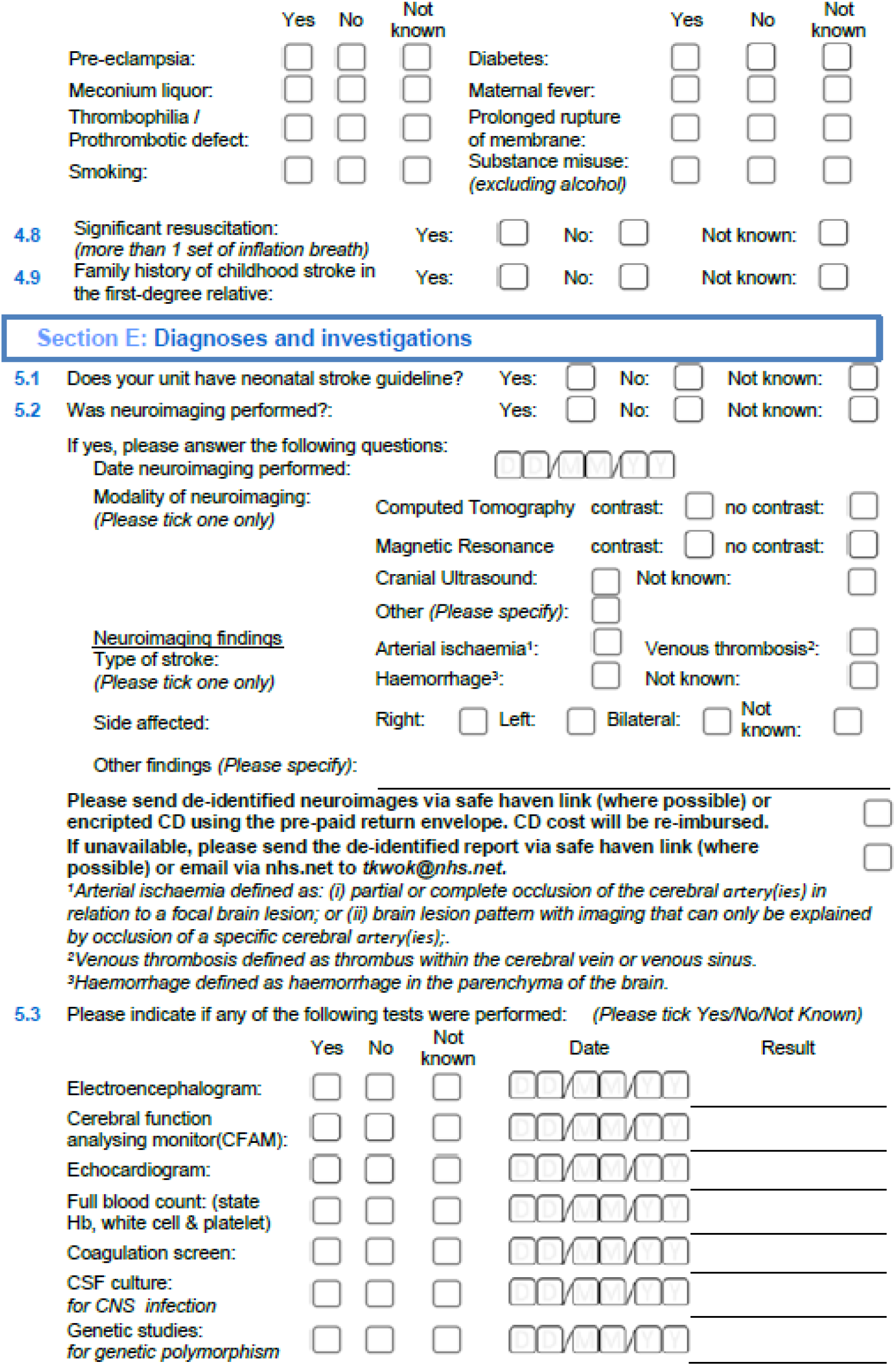
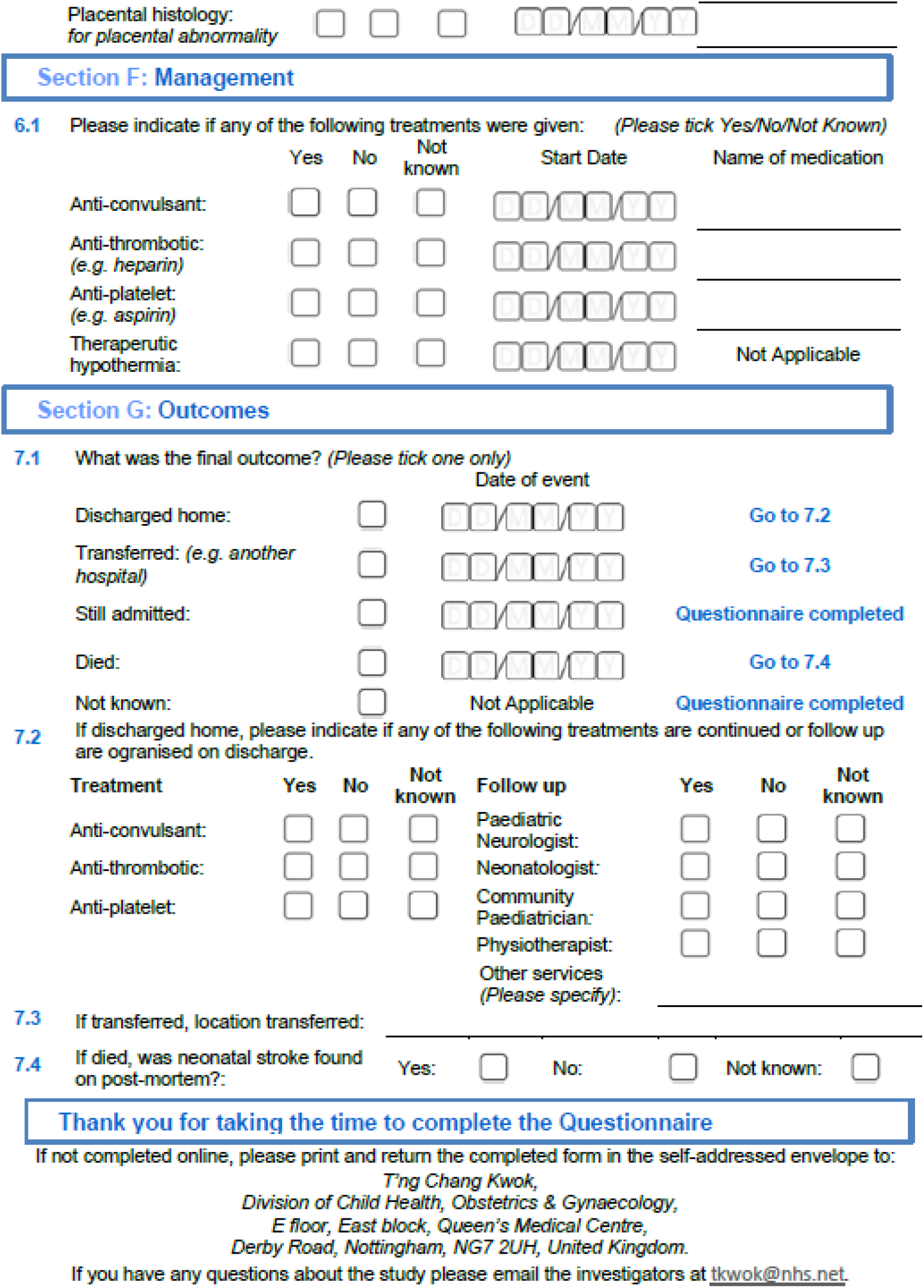

### Appendix 2: Two-year follow-up questionnaire

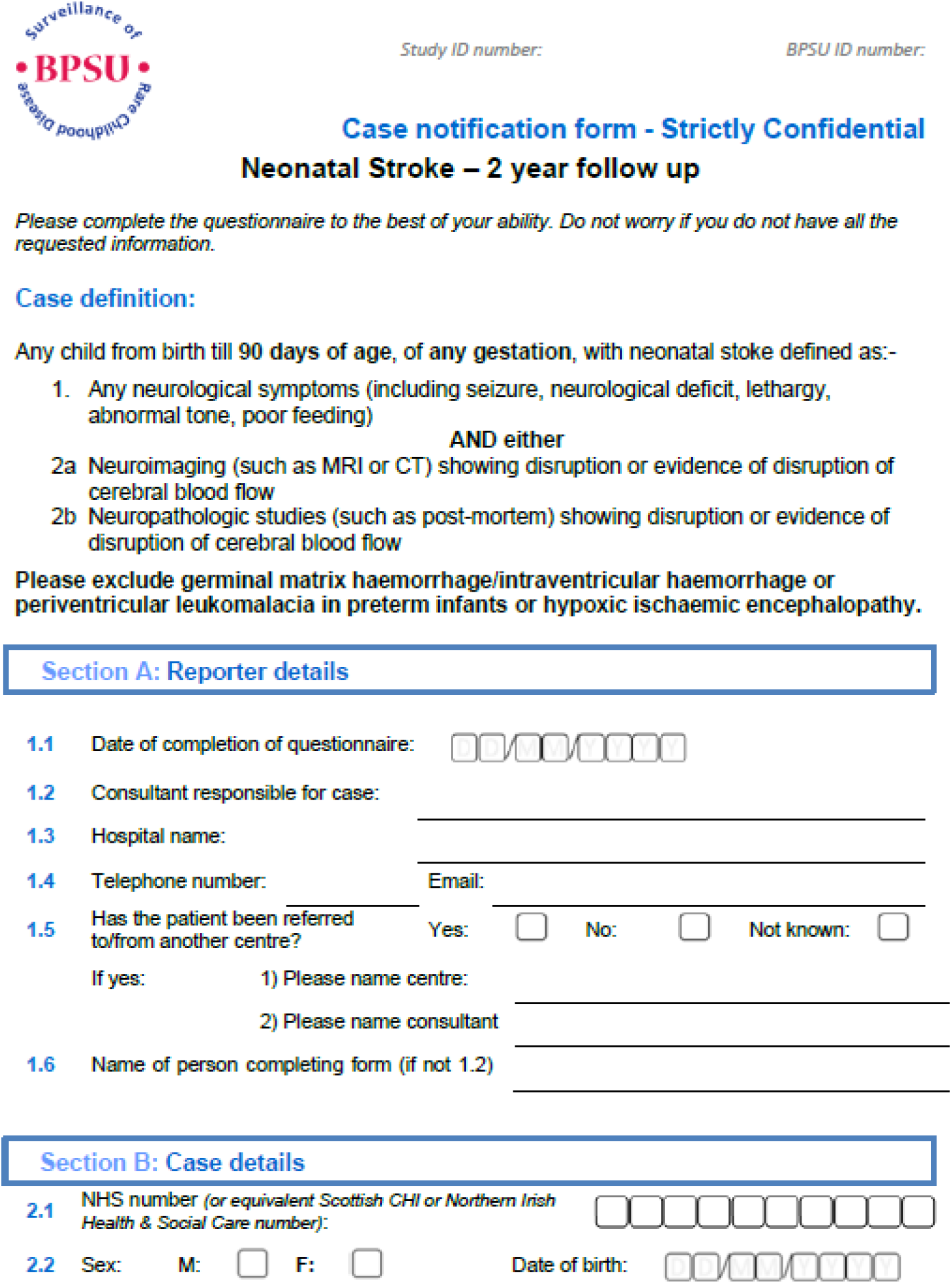
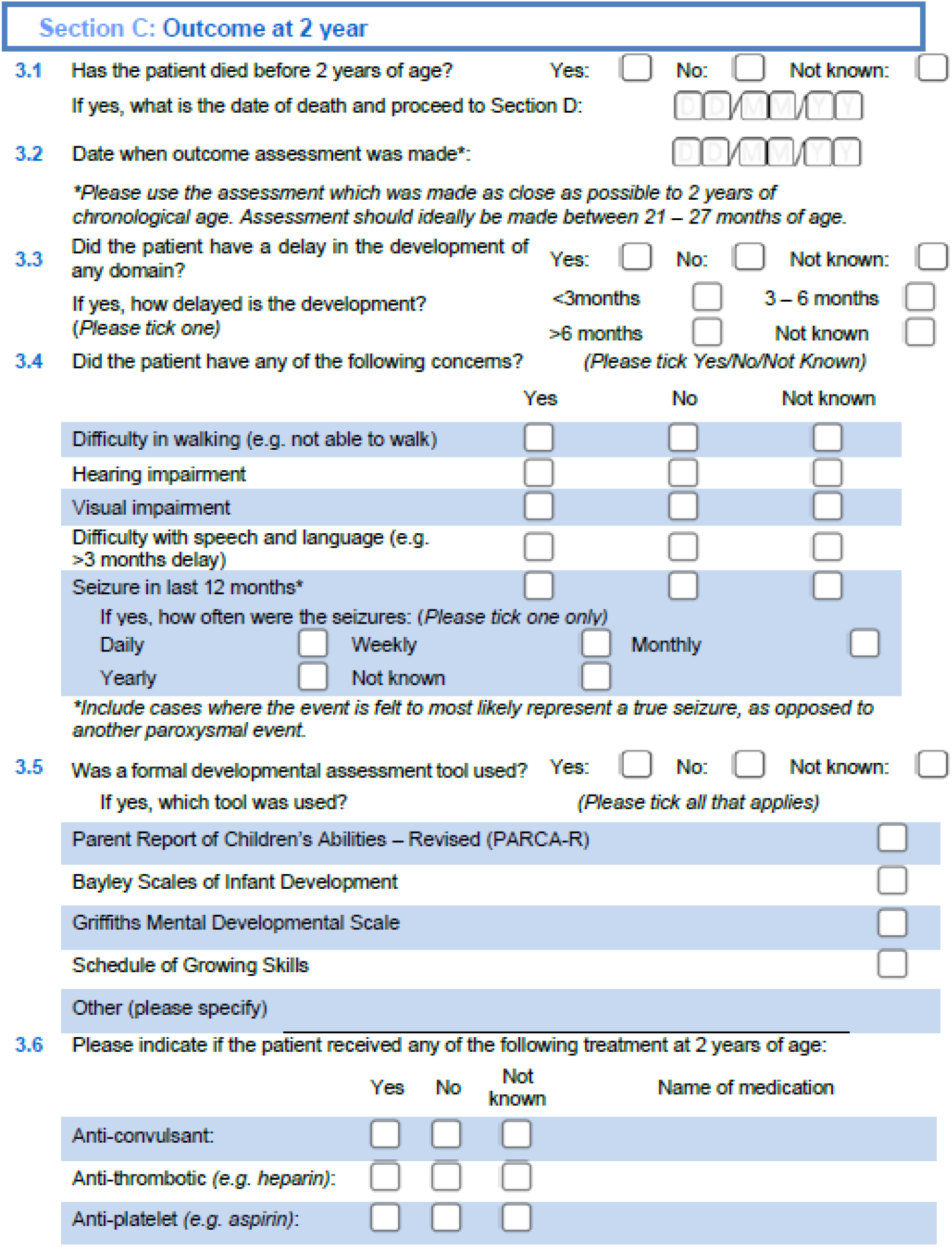
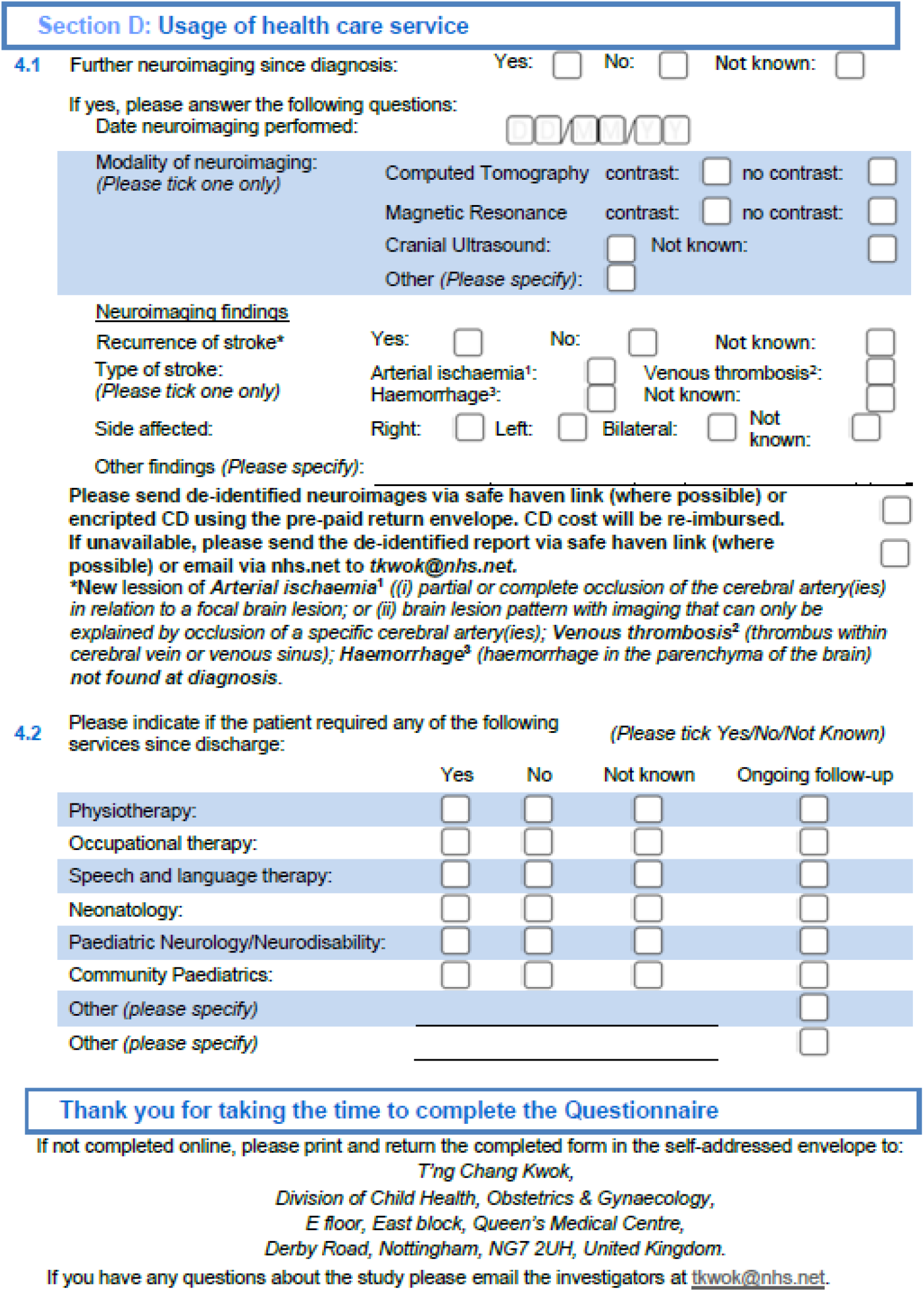

### Appendix 3: Initial questionnaire (Northern Ireland)

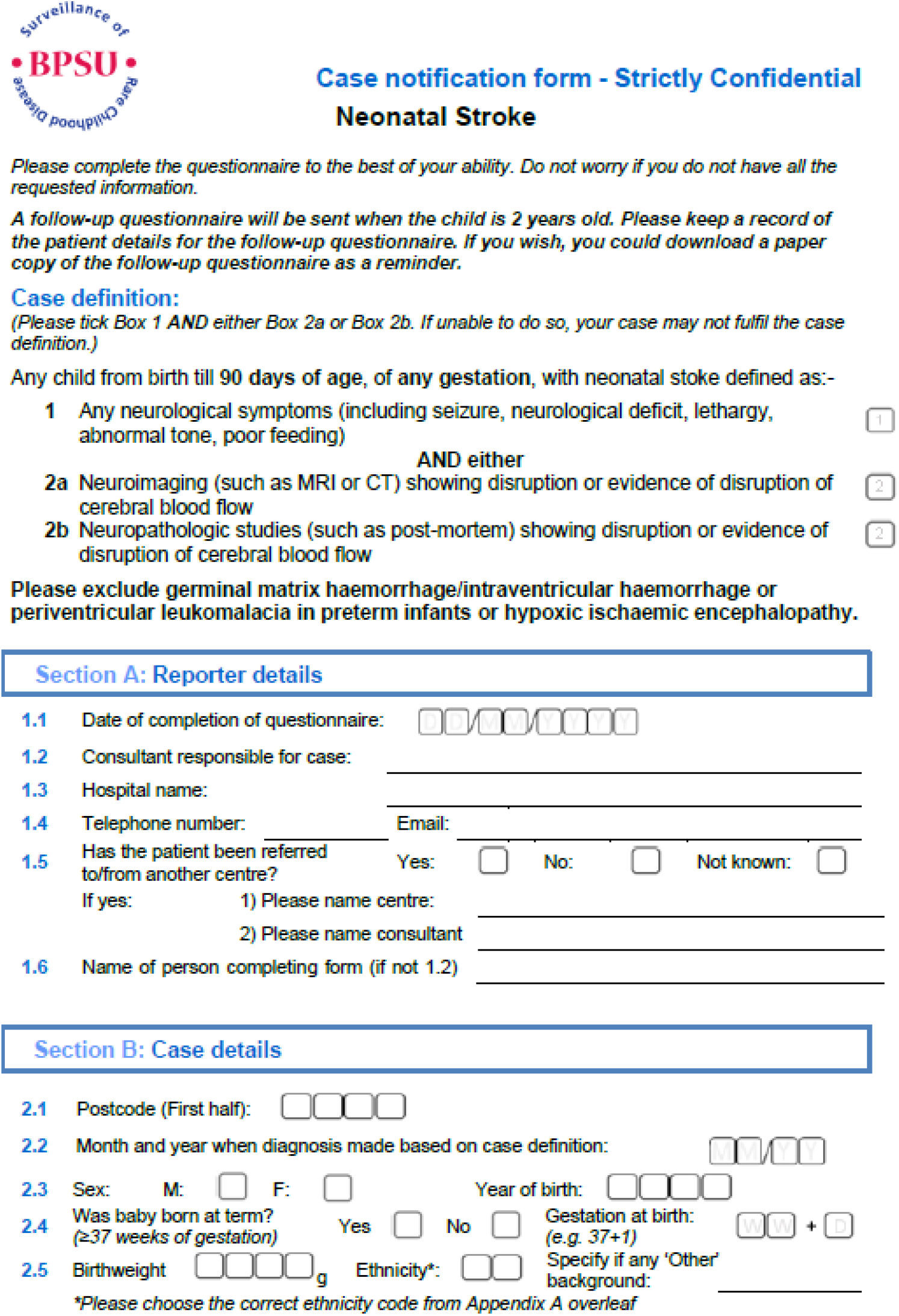
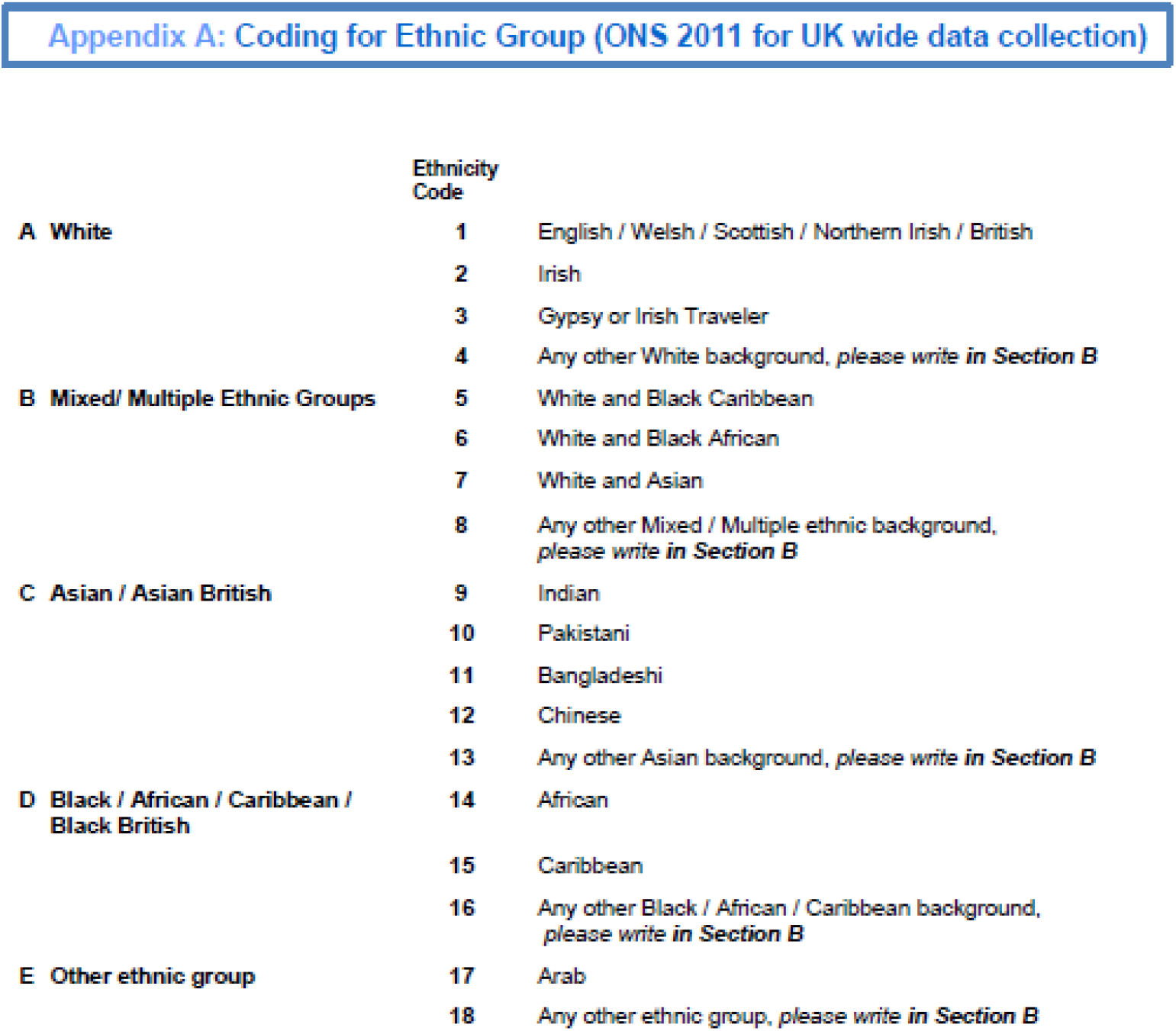
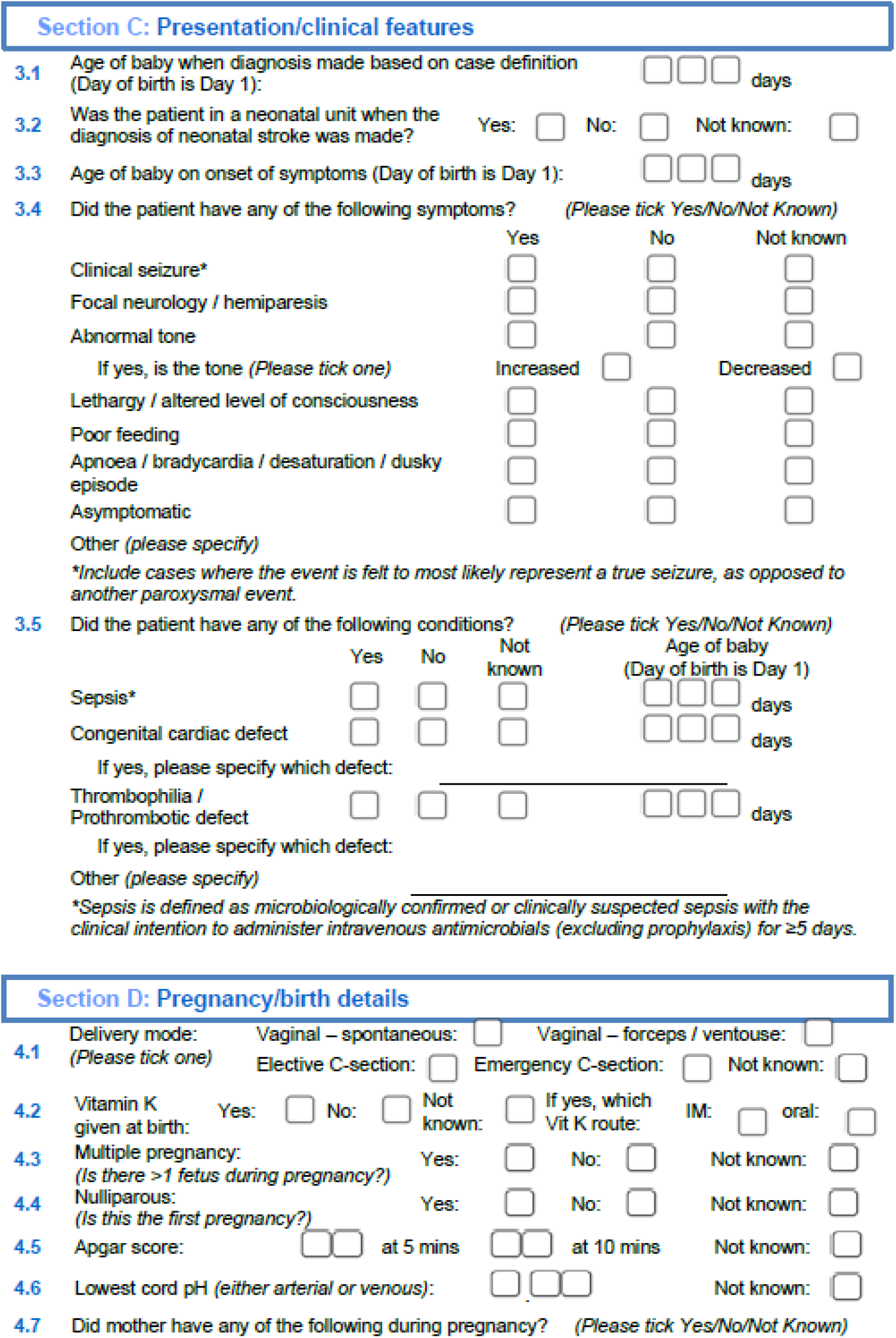
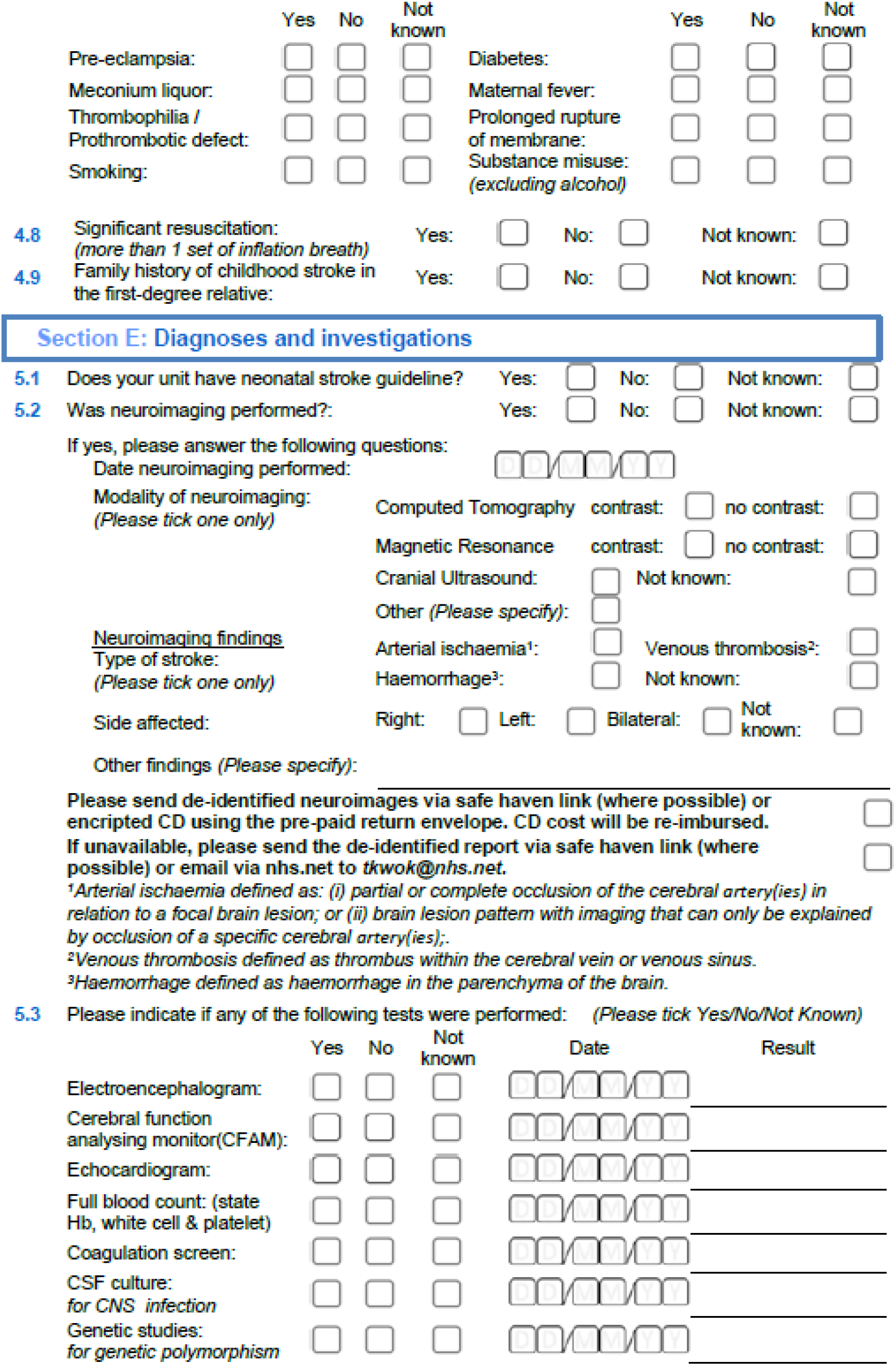
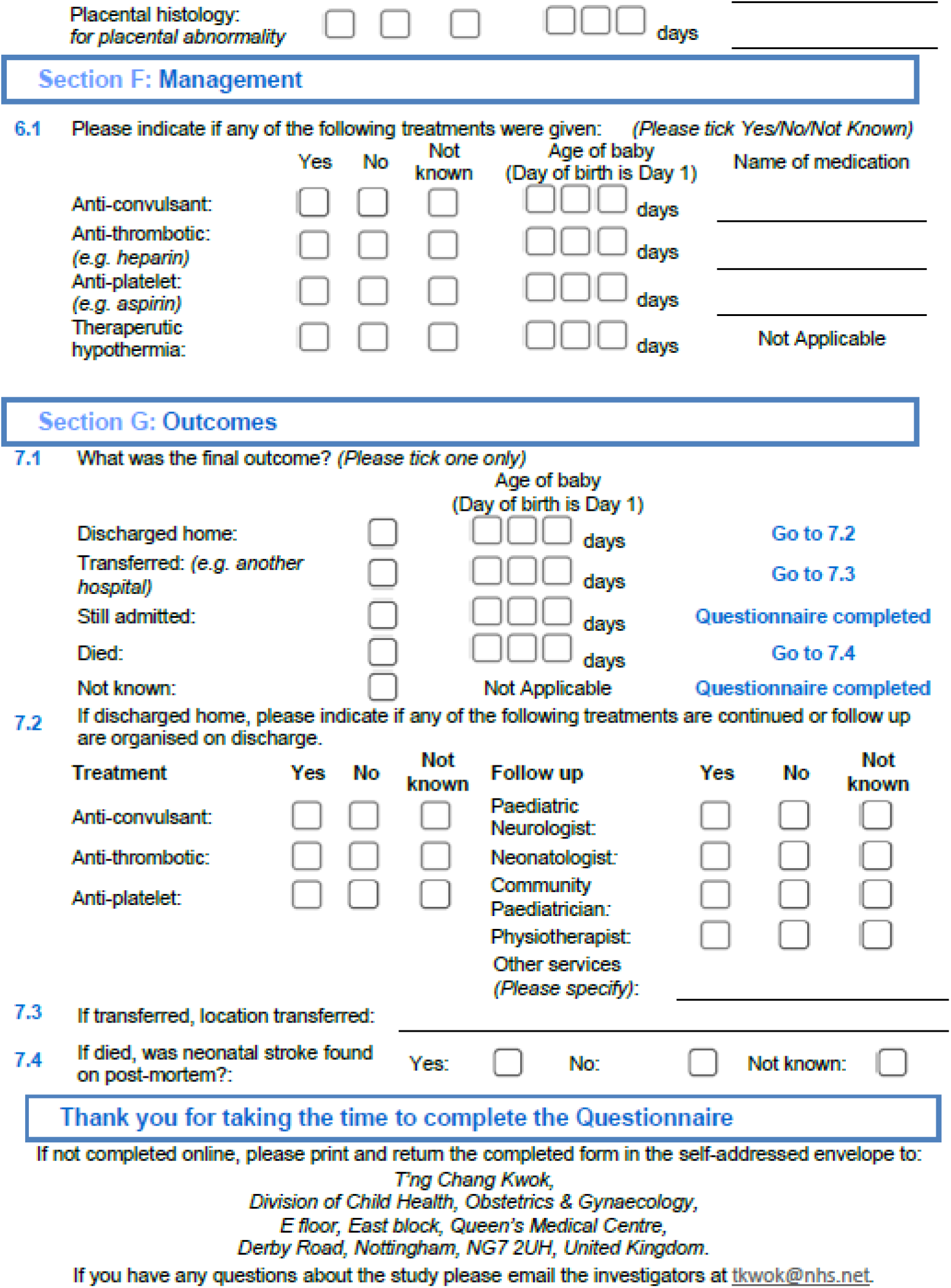

### Appendix 4: Two-year follow-up questionnaire (Northern Ireland)

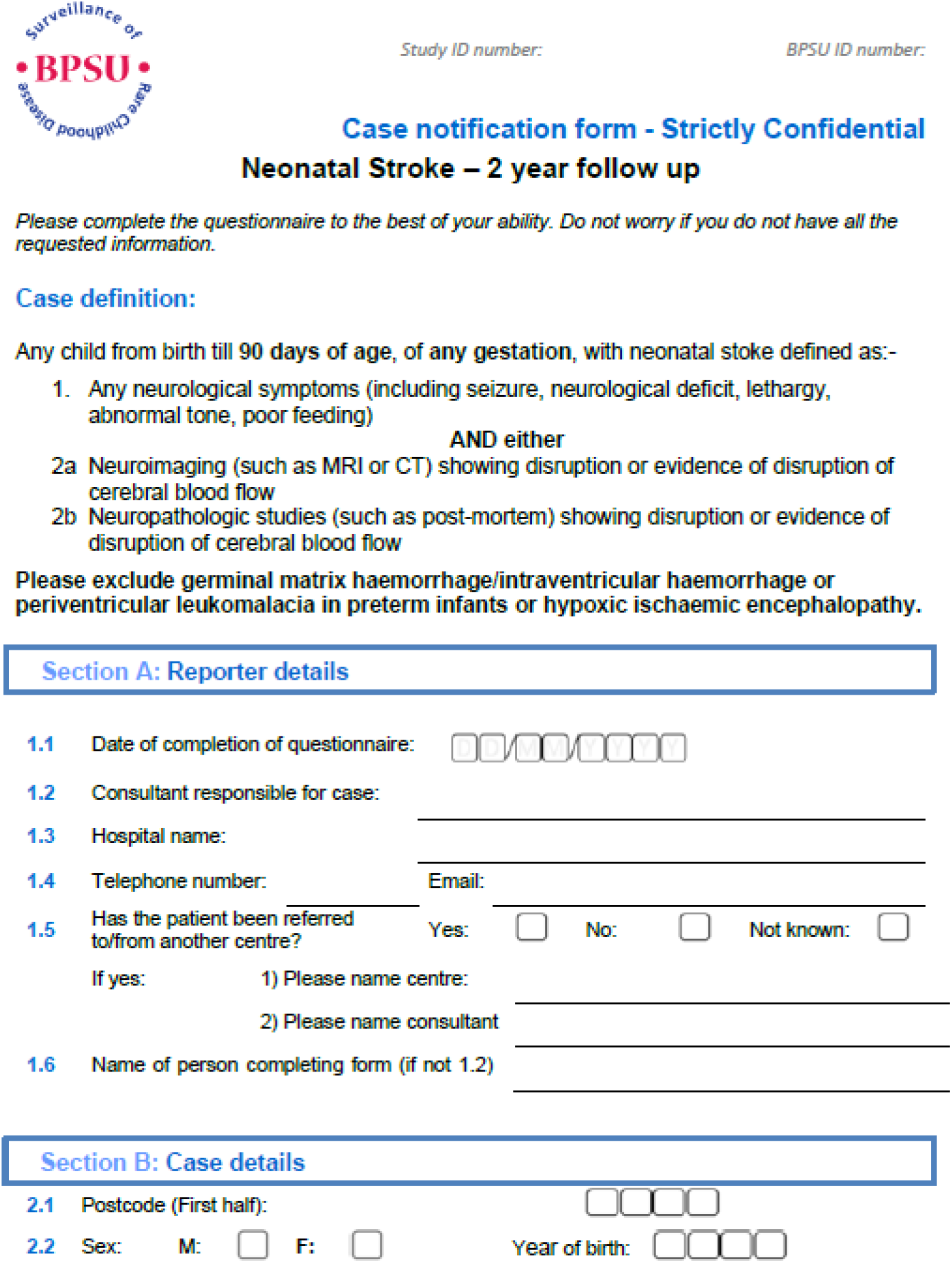
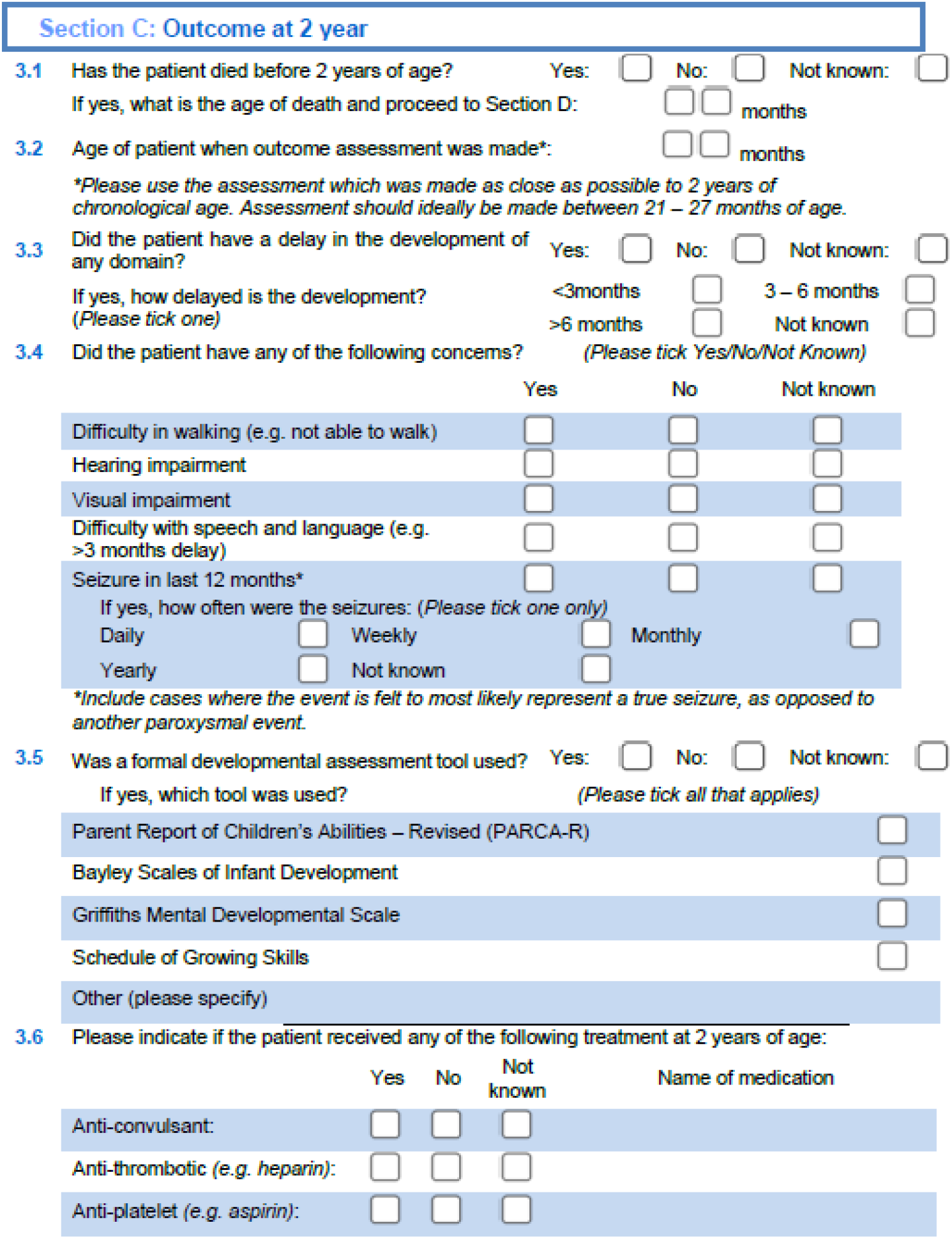
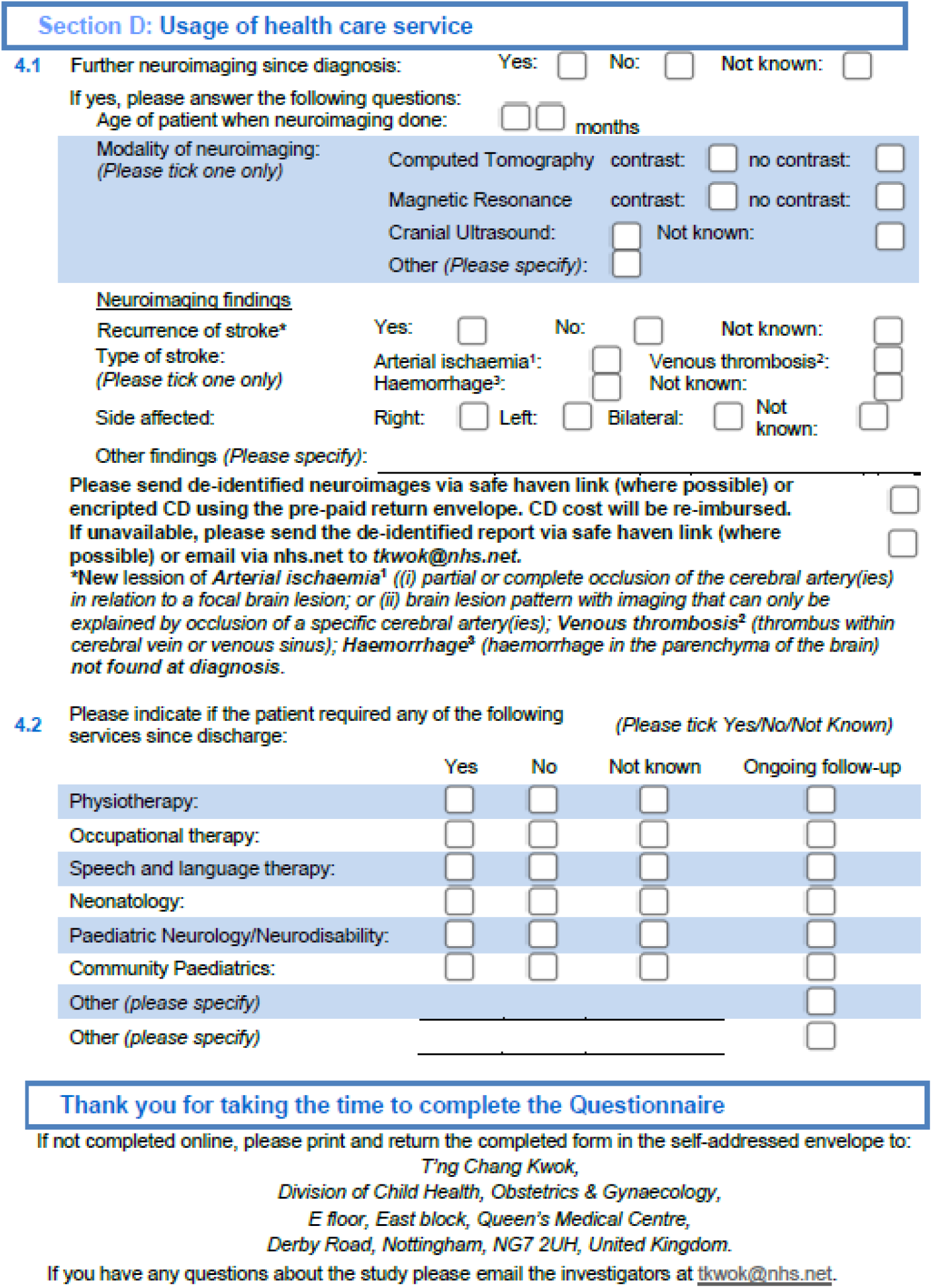

## References

1. Dunbar M, Kirton A. Perinatal stroke: mechanisms, management, and outcomes of early cerebrovascular brain injury. Lancet Child & Adolescent Health. 2018; 2:666–676.

2. Kirton A, Deveber G. Life after perinatal stroke. Stroke. 2013; 44:3265–3271.

3. Kirton A. Advancing non-invasive neuromodulation clinical trials in children: Lessons from perinatal stroke. Eur J Paediatr Neurol. 2017; 21:75–103.

4. Lai PT, Reilly JS. Language and affective facial expression in children with perinatal stroke. Brain Lang. 2015; 147:85–95.

5. Reilly JS, Wasserman S, Appelbaum M. Later language development in narratives in children with perinatal stroke. Dev Sci. 2013; 16:67–83.

6. Suppiej A, Mastrangelo M, Mastella L, Accorsi P, Grazian L, Casara G, et al. Pediatric epilepsy following neonatal seizures symptomatic of stroke. Brain Dev. 2016; 38:27–31.

7. Gardner MA, Hills NK, Sidney S, Johnston SC, Fullerton HJ. The 5-year direct medical cost of neonatal and childhood stroke in a population-based cohort. Neurology. 2010; 74:372–378.

8. Shepherd E, Salam RA, Middleton P, Han S, Makrides M, McIntyre S, et al. Neonatal interventions for preventing cerebral palsy: an overview of Cochrane Systematic Reviews. Cochrane Database Syst Rev. 2018; 6:CD012409.

9. Bemister TB, Brooks BL, Dyck RH, Kirton A. Predictors of caregiver depression and family functioning after perinatal stroke. BMC Pediatr. 2015; 15:75.

10. Bemister TB, Brooks BL, Dyck RH, Kirton A. Parent and family impact of raising a child with perinatal stroke. BMC Pediatr. 2014; 14:182.

11. Hart AR, Connolly DJA, Singh R. Perinatal arterial ischaemic stroke in term babies. Paediatrics and Child Health. 2018; 28:417–423.

12. Walther M, Juenger H, Kuhnke N, Wilke M, Brodbeck V, Berweck S, et al. Motor cortex plasticity in ischemic perinatal stroke: a transcranial magnetic stimulation and functional MRI study. Pediatr Neurol. 2009; 41:171–178

13. Rutherford MA, Ramenghi LA, Cowan FM. Neonatal stroke. Arch Dis Child Fetal Neonatal Ed. 2012; 97:F377–384.

14. deVeber GA, Kirton A, Booth FA, Yager JY, Wirrell EC, Wood E, et al. Epidemiology and Outcomes of Arterial Ischemic Stroke in Children: The Canadian Pediatric Ischemic Stroke Registry. Pediatr Neurol. 2017; 69:58–70.

15. Kirkham F, Williams A, Aylett S, Ganesan V. Cerebrovascular disease/stroke and like illness. British Paediatric Surveillance Unit – Annual Report 2002-2003. 2003; 10. https://www.rcpch.ac.uk/sites/default/files/2018-06/17th_annual_report.pdf (accessed on 02/02/2022).

16. Agrawal N, Johnston SC, Wu YW, Sidney S, Fullerton HJ. Imaging data reveal a higher pediatric stroke incidence than prior US estimates. Stroke. 2009; 40:3415–3421.

17. Armstrong-Wells J, Johnston SC, Wu YW, Sidney S, Fullerton HJ. Prevalence and predictors of perinatal hemorrhagic stroke: results from the kaiser pediatric stroke study. Pediatrics. 2009; 123:823–828.

18. Cole L, Dewey D, Letourneau N, Kaplan BJ, Chaput K, Gallagher C, et al. Clinical Characteristics, Risk Factors, and Outcomes Associated With Neonatal Hemorrhagic Stroke: A Population-Based Case-Control Study. JAMA Pediatr. 2017; 171:230–238.

19. Ferriero DM, Fullerton HJ, Bernard TJ, Billinghurst L, Daniels SR, DeBaun MR, et al. Management of Stroke in Neonates and Children: A Scientific Statement From the American Heart Association/American Stroke Association. Stroke. 2019; 50:E51–E96.

20. Knowles RL, Smith A, Lynn R, Rahi JS, Unit BPS. Using multiple sources to improve and measure case ascertainment in surveillance studies: 20 years of the British Paediatric Surveillance Unit. J Public Health (Oxf). 2006; 28:157–165.

21. Raju TN, Nelson KB, Ferriero D, Lynch JK, Participants N-NPSW. Ischemic perinatal stroke: summary of a workshop sponsored by the National Institute of Child Health and Human Development and the National Institute of Neurological Disorders and Stroke. Pediatrics. 2007; 120:609–616.

22. Sébire G, Fullerton H, Riou E, deVeber G. Toward the definition of cerebral arteriopathies of childhood. Curr Opin Pediatr. 2004; 16:617–622.

23. Govaert P, Ramenghi L, Taal R, de Vries L, deVeber G. Diagnosis of perinatal stroke I: definitions, differential diagnosis and registration. Acta Paediatrica. 2009; 98:1556–1567.

24. Gale C, Statnikov Y, Jawad S, Uthaya SN, Modi N, group BIew. Neonatal brain injuries in England: population-based incidence derived from routinely recorded clinical data held in the National Neonatal Research Database. Arch Dis Child Fetal Neonatal Ed. 2018; 103:F301–F306.

25. Medical Research Council. MRC ethics series Good research practice: Principles and guidelines. July 2012. https://www.ukri.org/wp-content/uploads/2021/08/MRC-0208212-Good-research-practice_2014.pdf (accessed on 02/02/2022).

26. Office for National Statistics – Births, 2001: Summary of key live birth statistics. https://webarchive.nationalarchives.gov.uk/ukgwa/20110116054739/ http://www.statistics.gov.uk/STATBASE/xsdataset.asp?vlnk=5672%26More=Y (accessed on 02/02/2022).

27. Gale C, Jeyakumaran D, Ougham K, Jawad S, Uthaya S, Modi N. Brain injury occurring during or soon after birth: annual incidence and rates of brain injuries to monitor progress against the national maternity ambition 2016 and 2017 data. London: National Data Analysis Unit, Imperial College London 2019.

28. Dunbar M, Mineyko A, Hill M, Hodge J, Floer A, Kirton A. Population Based Birth Prevalence of Disease-Specific Perinatal Stroke. Pediatrics. 2020; 146.

29. McLeish J, Alderdice F, Robberts H, Cole C, Dorling J, Gale C, et al. Challenges of a simplified opt-out consent process in a neonatal randomised controlled trial: qualitative study of parents’ and health professionals’ views and experiences. Arch Dis Child Fetal Neonatal Ed. 2021; 106:244–250.

30. NHS Health Research Authority. FAQs about the law. https://s3.eu-west-2.amazonaws.com/www.hra.nhs.uk/media/documents/cag-frequently-asked-questions-1.pdf (accessed on 02/02/2022).

31. Dunbar M, Kirton A. Perinatal Stroke. Semin Pediatr Neurol. 2019; 32:100767.

